# Ozone therapy for patients with COVID-19 pneumonia: preliminary report of a prospective case-control study

**DOI:** 10.1101/2020.06.03.20117994

**Authors:** Alberto Hernández, Montserrat Viñals, Asunción Pablos, Francisco Vilás, Peter J Papadakos, Duminda Wijeysundera, Sergio D. Bergese, Marc Vives

## Abstract

**Background:** There is still no specific treatment strategies for COVID-19 other than supportive management.

**Design:** A prospective case-control study determined by admittance to the hospital based on bed availability.

**Participants:** Eighteen patients with COVID-19 infection (laboratory confirmed) severe pneumonia admitted to hospital between 20th March and 19th April 2020. Patients admitted to the hospital during the study period were assigned to different beds based on bed availability. Depending on the bed the patient was admitted, the treatment was ozone autohemotherapy or standard treatment. Patients in the case group received ozonated blood twice daily starting on the day of admission for a median of four days. Each treatment involved administration of 200 mL autologous whole blood enriched with 200 mL of oxygen-ozone mixture with a 40 μg/mL ozone concentration.

**Main Outcomes:** The primary outcome was time from hospital admission to clinical improvement.

**Results:** Nine patients (50%) received ozonated autohemotherapy beginning on the day of admission. Ozonated autohemotherapy was associated with shorter time to clinical improvement (median [IQR]), 7 days [6-10] vs 28 days [8-31], p=0.04) and better outcomes at 14-days (88.8% vs 33.3%, p=0.01). In risk-adjusted analyses, ozonated autohemotherapy was associated with a shorter mean time to clinical improvement (−11.3 days, p=0.04, 95% CI -22.25 to -0.42).

**Conclusion:** Ozonated autohemotherapy was associated with a significantly shorter time to clinical improvement in this prospective case-control study. Given the small sample size and study design, these results require evaluation in larger randomized controlled trials.

## Introduction

The COVID-19 pandemic has led to more than 28.7 million cases and 920.847 deaths globally as of September 2020. ^1^ About 15% of infected adults develop severe pneumonia requiring supplemental oxygen, and an additional 5% progress to acute respiratory distress syndrome (ARDS) requiring mechanical ventilation often for several weeks. ^2,3^

Supportive measures remain the cornerstone for treating COVID-19 in the absence of specific therapies. The potential biological benefits of ozonated autohemotherapy include reduced tissue hypoxia, decreased hypercoagulability, modulated immune function with inhibition of inflammatory mediators, improved phagocytic function, and impaired viral replication. ^4-13^

Ozone might improve blood circulation and oxygen delivery to ischemic tissue ^4-7^ as a result of the concerted effect of nitric oxide, ^8^ increase intra-erythrocytic 2,3-DPG level, ^9^ and increase of some prostacyclins such as PGI2. ^10^ These effects can help to decrease the hypercoagulation that has been observed in COVID-19 patients. ^11^ Another important role played by ozone in COVID-19 is its immunomodulatory effects. The inflammatory response is a hallmark of severe infection and cytokine modulation is key to avoid patient deterioration. Ozone is able to modulate and control cytokines releasing anti-inflammatory cytokines and reducing activity of pro-inflammatory such as IL-1, IL-6 and TNF-α counteracting the state of hyperinflammation seen in COVID patients, but in addition, ozone has potent anti-inflammatory properties through the modulation of the NLRP3 inflammasome which is recognized to play a crucial role in the initiation and continuance of inflammation in various diseases. ^12^ Ozone may also modulate the accumulation of neutrophils locally, the expression of IL-6, TNF-α, and albumin modified by ischemia in the kidneys, as well as increase local antioxidant capacity. ^13^

Ozone therapy is the administration of a mixture of gas of 97% oxygen and 3% ozone generated from a medical ozone generator. Ozone is a molecule which consists of three oxygen atoms all sharing the same electrons. Because there just are not enough electrons to go around, ozone is a relatively unstable molecule. This instability is why it is such a powerful biological stimulant. ^14^ Ozone therapy can be administered systemically by adding it to a sample of a patient’s own blood sample and then reinfusing it, in what is termed ‘ozonated autohemotherapy’.

Ozone is a naturally occurring gas produced from oxygen atoms. Single oxygen atoms cannot endure alone without being regrouped into di-atomic oxygen molecules. In this recombination phase, some atoms will transform into loosely bound tri-atomic oxygen. This novel trioxygen molecule is called ozone which is found in the stratosphere where absorbs various ultraviolet radiation to protect us. Its molecular weight is of 48 g/mol with a solubility in water of 0.57 g/L at a temperature of 20 °C, (about ten-fold higher than oxygen). Consequently, the great solubility of ozone in water allows its immediate reaction with any soluble compounds and biomolecules present in biological fluids.

Ozone is generated by medical devices for medical purposes. Medical ozone is obtained from pure oxygen by passing it through a high voltage gradient (5-13 KV). This yields a gas mixture consisting of 97% oxygen and no more than 3% ozone. Thermodynamically is unstable and spontaneously reverts back into oxygen. Concentrations ranging from 10-70 μg/ml are commonly used for medical purposes. There are multiples routes for medical ozone administration. Inhalation route may be toxic to the pulmonary system and other organs. However, ozonated autohemotherapy has been shown to be safe in multiple randomized clinical trials, observational studies and meta-analyses. ^15^ The incidence of side effects of ozone therapy is very low (estimated at 0.0007%), and typically manifests itself as euphoria, nausea, headaches and fatigue. ^16^ In general, it is a very safe therapy when administered correctly, with the recommended dose. Complications like air embolism have been described ^17^ but are caused by incorrect administration practices and by using non-certified equipment.

Several countries including Spain, Italy, Greece, Cuba, Russia, Portugal and Turkey have incorporated ozone therapy in medical practice for other indications. ^18^

The pathogenesis of the virus is variable and not fully understood. It predominantly involves the lungs where diffuse alveolar damage with involvement of the microcirculation leads to marked hypoxia. ^19, 20^ A dysregulation of the immune response is present and lymphocytopenia is a hallmark in the vast majority of these patients. ^21^ Innate immunity and coagulation pathways are intricately linked. ^22^ COVID-19–associated macrophage activation, hyperferritinemia, cytokine storm, release of pathogen-associated molecular patterns and damage-associated molecular proteins can result in release of tissue factor and activation of coagulation factors that create a predisposition to hypercoagulability. ^22^

Others have reported, as case reports, the use of ozonated autohemotherapy in patients with severe COVID-19 pneumonia, however, they had limitations. ^23-25^ A retrospective case-control study, on 60 patients with mild to moderate COVID-19 pneumonia has been recently published. ^26^

We, therefore, conducted a prospective case-control study determined by admittance to the hospital based on bed availability to determine if ozonated autohemotherapy was associated with a shorter time to clinical improvement in patients with severe COVID-19 pneumonia.

## Materials and methods

### Study Design

This prospective case-control study was performed at the Policlinica Ibiza Hospital in Spain. It was conducted in compliance with the Declaration of Helsinki and approved by a multidisciplinary human research ethics committee (HREC) at the institution. Each participant gave written informed consent for administration of any interventions, collection of relevant clinical data and ascertainment of outcomes. The study consisted of all adults (aged ≥18 years) who were admitted to the hospital with a diagnosis of severe COVID-19 pneumonia between 20^th^ March to 19^th^ April 2020. All included patients met the following criteria: confirmed COVID-19 infection (diagnosed by nasopharyngeal swab performed on admission); pneumonia with baseline chest X-ray abnormalities; oxygen saturation <94% on room air, and tachypnea with respiratory rate exceeding 30 per minute.

Patients admitted to the hospital during the study period were assigned to different beds based on bed availability. Depending on the bed the patient was admitted, the treatment was ozone autohemotherapy or standard treatment.

### Standard Clinical Care

Treatment for all COVID-19 pneumonia patients included supplemental oxygen therapy, hydroxychloroquine, lopinavir/ritonavir, corticosteroids, and antibiotics (including azithromycin) at the discretion of the individual patient’s attending physician. <u>Drugs dosage were the standard dose: ceftriaxone 2gr q24h for 5 days, levofloxacino 500mg q12h, hydroxicloroquine 400mg q24h for 4 days, dexamethasone 6mg q24h for 10 days or methylprednisolone 40mg q12h and azithromycin 500mg q24h for 3 days. Neither remdesivir nor tocilizumab were given to any patient. Enoxaparin 1mg/kg SC q12h was used as therapeutic anticoagulation dose.</u> Decisions on endotracheal intubation, mechanical ventilation and critical care unit admission were made following clinical standards and at the discretion of the patient’s attending physician.

### Ozonated Autohemotherapy Intervention

Ozonated blood was given twice a day for 5 consecutive days. Ozonated autohemotherapy involved intravenous infusion of ozonated autologous whole blood. Initially, 200 mL of autologous whole blood was drawn from the patient’s antecubital vein into a standard plastic disposable blood collection bag (certified SANO_3_ bag) containing 35 mL of anticoagulant citrate dextrose solution (ACD-A). The blood was then enriched with 200 mL of gas mixture oxygen-ozone with an ozone concentration at 40 μg/mL obtained by *Ozonobaric P Sedecal*, an ozone generator with CE0120 certificate type IIb. The ozonized blood was then re-infused into the same vein over approximately 10-15 minutes. ^26^

### Outcomes

#### Primary outcome

The primary clinical outcome was time to clinical improvement during hospital admission.

#### Clinical evaluation

Clinical improvement was defined as a two-point reduction (relative to the patient’s status on hospital admission) on a six-point ordinal scale, or discharge alive from the hospital, whichever came first. The six-point scale was as follows: death (6 points); extracorporeal membrane oxygenation or mechanical ventilation <u>requiring intubation</u> (5 points); noninvasive ventilation or high-flow oxygen therapy (4 points); oxygen therapy without need for high-flow oxygen or non-invasive ventilation (3 points); hospital admission without need for oxygen therapy (2 points); and discharged from hospital or reached discharge criteria (1 point). Discharge criteria were as evidence of clinical recovery (normalization of pyrexia, respiratory rate <24 per minute, oxygen saturation >94% on room air, and absence of cough) for at least 72 hours.

This six-point scale and definition of clinical improvement (i.e., two-point improvement in scale) has been used in prior research on intervention for relating to COVID-19 infection. ^28^ Personnel ascertaining outcomes were not blinded to whether patients received usual care versus ozonated autohemotherapy.

#### Secondary outcomes

Secondary outcomes were clinical improvement as measured at the 7th, 14th and 28th days after hospital admission. Time to a two-fold decrease in concentrations of C-reactive protein, ferritin, D-dimer and lactate dehydrogenase were also daily measured. <u>Other secondary outcomes were the following: ventilator-free days at day 28, intubation rate, hospital length of stay, in-hospital and 28-days mortality and time (days) to PCR COVID-19 negative.</u> Follow-up ceased at the point of hospital discharge, patient death, or 31 days following hospital admission, which ever came first.

### Statistical Analysis

All analyses were performed using STATA version 13.0 (StataCorp. 2013. Stata Statistical Software: Release 13. College Station, TX: StataCorp LP). Statistical significance was defined by a 2-sided P-value less than 0.05. The Shapiro-Wilk test was used to determine whether variables were normally distributed. Unadjusted differences between treatment and control arms were then calculated using the two-sample t-test (normally distributed continuous variables), Mann-Whitney U-test (continuous variables with evidence of non-normal distributions) and Fisher’s exact test (categorical variables). Unadjusted times to clinical improvement were compared between the two study arms using Kaplan-Meier survival curves and the log-rank test. Patients were censored at the point of hospital discharge, death or 31 days following hospital admission, whichever came first. The adjusted association between ozonated autohemotherapy and mean time to clinical improvement was estimated using a multivariable linear regression model that adjusted for age, sex, and baseline quick Sequential Organ Failure Assessment (SOFA) score. These covariates were pre-specified on the basis of their clinical significance. Patients who had not achieved clinical improvement within the follow-up period were assigned a time value of 31 days. All patients admitted to the study site within a pragmatic one-month period were included in the study cohort.

## Results

The cohort included 18 patients. The mean age was 68 years old (SD 15 years) and 72.2% (n=13) were male. The baseline characteristics of these patients are presented in Table 1. In total, 9 patients (50%) received ozonated autohemotherapy. The baseline characteristics of the two study arms were qualitatively similar, aside from age (mean age was higher in the usual care arm), weight (mean weight was higher in the usual care arm), and body mass index (mean value was higher in the usual care only arm).

**Table 1.** Baseline characteristics

|  | **Ozonated**<br>**Autohemotherapy**<br>**(n=9)** | **Usual Clinical**<br>**Care**<br>**(n= 9)** | **p-value** |
| --- | --- | --- | --- |
| Age, mean (SD), years | 64 (11) | 71 (18) | 0.35 |
| Male sex, n (%) | 7 (78%) | 6 (67%) | 1 |
| Weight, mean (SD), kg | 74 (17) | 85 (23) | 0.25 |
| Height, mean (SD), cm | 167 (10) | 170 (7) | 0.48 |
| Body mass index, mean (SD), kg/m2 | 26.2 (4.5) | 29.5 (7.1) | 0.26 |
| Hypertension, n (%) | 4 (44%) | 6 (67%) | 0.34 |
| Diabetes mellitus, n (%) | 0 (0%) | 2 (22%) | 0.47 |
| Chronic pulmonary disease, n (%) | 2 (22%) | 1 (11%) | 1 |
| Chronic cardiac disease, n (%) | 1 (11%) | 2 (22%) | 1 |
| Previous stroke, n (%) | 0 (0%) | 0 (0%) | 1 |
| Baseline hemoglobin, mean (SD), mg/dL | 13 (2.1) | 13 (3.0) | 0.51 |
| Baseline Quick SOFA score of 2 or 3, n (%) | 1 (11%) | 1 (11%) | 1 |
| Baseline 6-point ordinal scale, median [IQR] | 3 [3-3] | 3 [2-3] | 0.60 |
| Baseline Lactate Dehydrogenase, mean (SD), U/L | 487 (168) | 506 (123) | 0.80 |
| Baseline C-reactive protein, median ([IQR]), mg/L | 2,9 [0,5-7,75] | 4,3 [1,8-8,9] | 0.50 |
| Baseline ferritin, median ([IQR], ug/L | 556 [226-1,171] | 290 [163-880] | 0.63 |
| Baseline D-dimer, median ([IQR], ng/mL | 943 [459-1,930] | 389 [215-468] | 0.16 |
| Baseline platelets, median ([IQR], $\times 10^9$ /L | 302 [263-408] | 180 [155-211] | 0.05 |
| Baseline SpO2/FiO2 ratio, median [IQR] | 350 [255-408] | 339 [261-452] | 0.96 |
| **Treatment** |  |  |  |
| Hydroxychloroquine, n (%) | 4 (44%) | 3 (33%) | 0.23 |
| Lopinavir/ritonavir, n (%) | 1 (11 %) | 2 (22%) | 1 |
| Corticosteroids, n (%) | 2 (22%) | 1 (11%) | 1 |
| Ceftriaxone, n (%) | 1 (11%) | 1 (11%) | 1 |
| Levofloxacin, n (%) | 2 (22%) | 2 (22%) | 1 |
| Azithromycin, n (%) | 8 (89%) | 7 (78%) | 1 |
| Therapeutic anticoagulation, n (%) | 0 (0 %) | 2 (22%) | 1 |

IQR: Interquartile range. SD: Standard difference. SOFA: Sequential Organ Failure Assessment.

### Primary outcome: Time to clinical improvement

Ozonated autohemotherapy was associated with a significantly lower time to clinical improvement (median [IQR]), 7 days [6-10] vs 28 days [8-31], p=0.04) (Figure 1 and Table 2). In unadjusted linear regression analyses, the mean time to clinical improvement was 12.4 days shorter in the ozonated autohemotherapy arm (−12.4 days; p=0.01; 95% CI -22.49 to -2.39). In adjusted linear regression analyses, the mean time to clinical improvement in the ozonated autohemotherapy arm was 11.3 days shorter (−11.3 days, p=0.04, 95% CI -22.25 to -0.42). We conducted a post-hoc sensitivity analysis that adjusted for age, quick SOFA and weight – all of which were baseline characteristics with qualitative differences between study arms. The adjusted difference in time to clinical improvement (−11.6 days, p=0.05, 95% CI -23.3 to 0.41) was qualitatively similar in this sensitivity analysis. Unadjusted times to clinical improvement using Kaplan-Meier survival curves and the log-rank test showed a significant difference between groups (Log Rank (Mantel-Cox) Chi-square 4,182. p=0,041) (Fig 1).

**Table 2.**
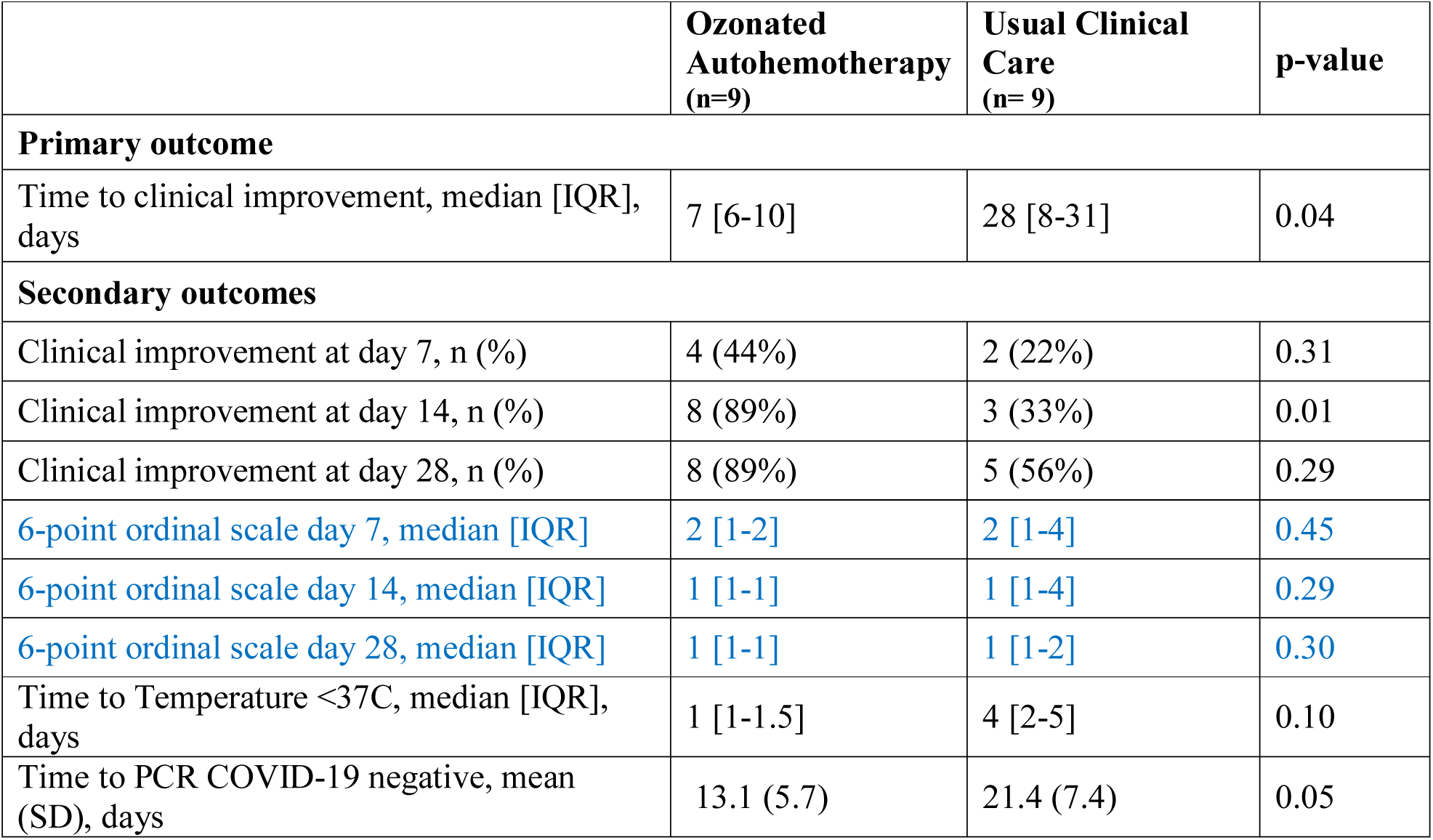
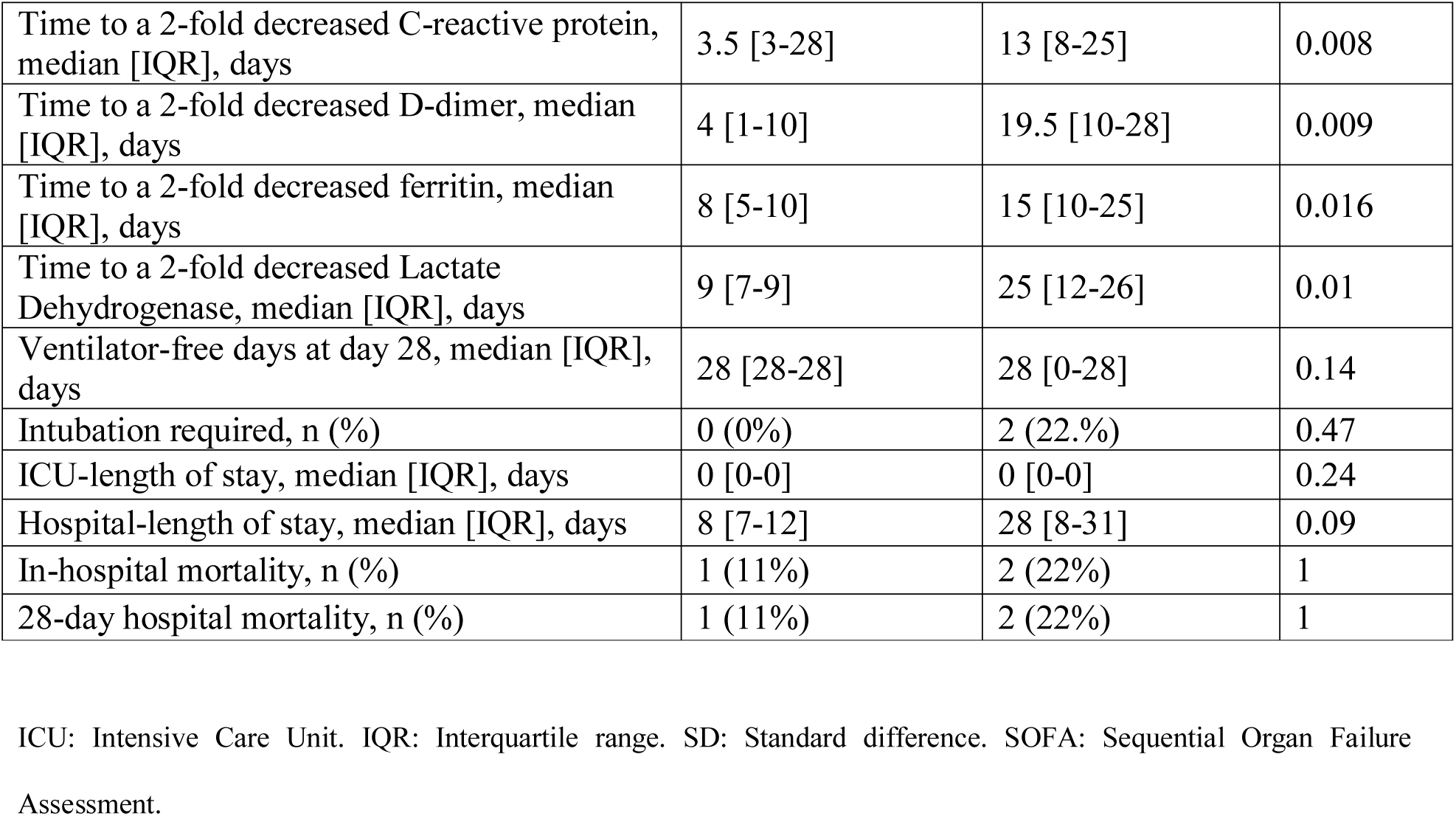
Outcomes

|  | **Ozonated**<br>**Autohemotherapy**<br>**(n=9)** | **Usual Clinical**<br>**Care**<br>**(n= 9)** | **p-value** |
| --- | --- | --- | --- |
| **Primary outcome** |  |  |  |
| Time to clinical improvement, median [IQR], days | 7 [6-10] | 28 [8-31] | 0.04 |
| **Secondary outcomes** |  |  |  |
| Clinical improvement at day 7, n (%) | 4 (44%) | 2 (22%) | 0.31 |
| Clinical improvement at day 14, n (%) | 8 (89%) | 3 (33%) | 0.01 |
| Clinical improvement at day 28, n (%) | 8 (89%) | 5 (56%) | 0.29 |
| 6-point ordinal scale day 7, median [IQR] | 2 [1-2] | 2 [1-4] | 0.45 |
| 6-point ordinal scale day 14, median [IQR] | 1 [1-1] | 1 [1-4] | 0.29 |
| 6-point ordinal scale day 28, median [IQR] | 1 [1-1] | 1 [1-2] | 0.30 |
| Time to Temperature <37C, median [IQR], days | 1 [1-1.5] | 4 [2-5] | 0.10 |
| Time to PCR COVID-19 negative, mean (SD), days | 13.1 (5.7) | 21.4 (7.4) | 0.05 |
| Time to a 2-fold decreased C-reactive protein, median [IQR], days | 3.5 [3-28] | 13 [8-25] | 0.008 |
| Time to a 2-fold decreased D-dimer, median [IQR], days | 4 [1-10] | 19.5 [10-28] | 0.009 |
| Time to a 2-fold decreased ferritin, median [IQR], days | 8 [5-10] | 15 [10-25] | 0.016 |
| Time to a 2-fold decreased Lactate Dehydrogenase, median [IQR], days | 9 [7-9] | 25 [12-26] | 0.01 |
| Ventilator-free days at day 28, median [IQR], days | 28 [28-28] | 28 [0-28] | 0.14 |
| Intubation required, n (%) | 0 (0%) | 2 (22.%) | 0.47 |
| ICU-length of stay, median [IQR], days | 0 [0-0] | 0 [0-0] | 0.24 |
| Hospital-length of stay, median [IQR], days | 8 [7-12] | 28 [8-31] | 0.09 |
| In-hospital mortality, n (%) | 1 (11%) | 2 (22%) | 1 |
| 28-day hospital mortality, n (%) | 1 (11%) | 2 (22%) | 1 |

ICU: Intensive Care Unit. IQR: Interquartile range. SD: Standard difference. SOFA: Sequential Organ Failure Assessment.

**Figure 1.**
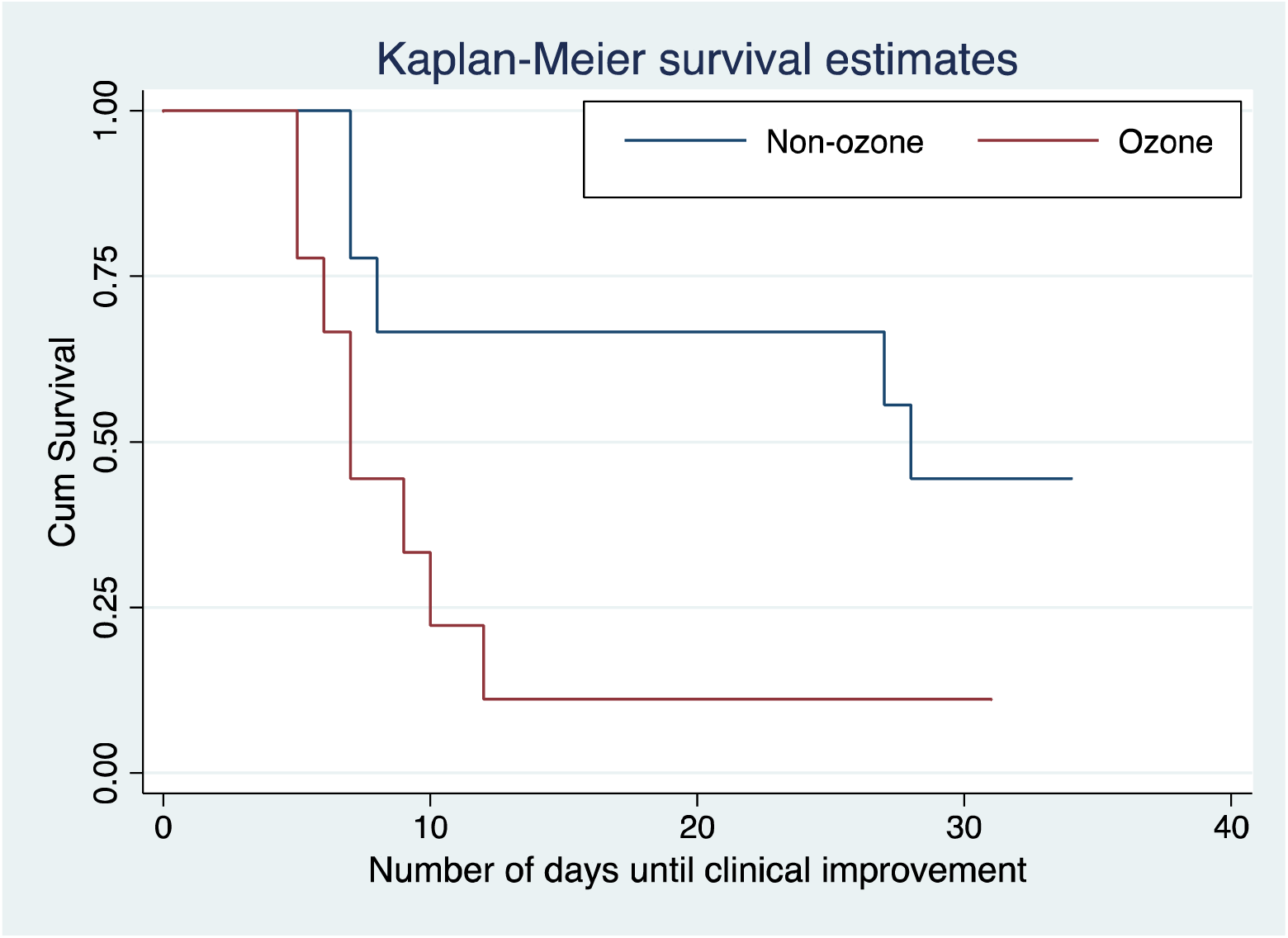
Kaplan-Meier survival curves

### Secondary Outcomes

Ozonated autohemotherapy was associated with clinical improvement at day 14 (88.8% vs 33.3%, p=0.01). Ozonated autohemotherapy was also associated with a shorter time to a two-fold decrease of C-reactive protein (3.5 days [3-28] vs 13 days [8-25], p=0.008), ferritin (8 days [5-10] vs 15 days [10-25], p=0.016), D-dimer (4 days [1-10] vs 19.5 days [10-28], p=0.009) and Lactate Dehydrogenase (9 days [7-9] vs 25 days [12-26], p=0.01). The mean time to negative PCR COVID-19 testing results was reduced [13.1 (SD 5.7) vs 21.4 (SD 7.4 days), p=0.05). There was no difference with respect to ventilator-free days at day 28 (median [IQR]), 28 days [28-28] vs 28 days [0-28], p=0.14) or 28-days mortality (11.1% vs 22.2%; p=1). No adverse events were observed or unintended effects in both groups. None of the patients in both groups were treated with non-invasive mechanical ventilation.

## Discussion

In this prospective case-control study of 18 patients with confirmed COVID-19 severe pneumonia, twice-daily ozonated autohemotherapy for 5 consecutive days was associated with a significant reduction in the time to clinical improvement. This case-control study provides novel new data pointing to the potential role of ozonated autohemotherapy for treatment of severe COVID-19 pneumonia.

Our findings are consistent with recent reviews describing the potential biologically plausible benefits associated with ozonated autohemotherapy for COVID-19 ^29-32^ and also consistent with a recently published retrospective case-control study. ^26^

Tascini et al. in their case-control study, ^26^ on 60 patients with mild to moderate COVID-19 pneumonia, treated both groups with best available therapy, showed an association between the use of blood ozonization and a significant decrease on the SIMEU clinical phenotype according to the Italian Society of Emergency and Urgency Medicine (2.87 ± 0.78 vs. 2.27 ± 0.83, p < 0.001) from baseline to discharge. Whereas in the control group there was no statistically significant difference. Furthermore, the clinical improvement associated with the use of O3 was greater compared to the control group (53% vs 33%). In the case group, only 7% of patients had a worse outcome, compared with 17% in the control group. As in our cohort, no adverse events associated to the treatment with ozonated blood were observed. Among the 30 patients treated with ozonated blood (cases group), 28 received three consecutive sessions, and 2 received two consecutive doses for 3 days. The dose used was 200 mL of gas mixture oxygen-ozone with an ozone concentration at 40μg/mL. In our study, the same dose was received. However, it was given twice a day during 5 days, instead of 3 sessions per day as they did. In our opinion, the primary endpoint in Tascini et al. study was somewhat confusing. There was a decrease on the SIMEU clinical phenotype from baseline to discharge and the clinical improvement associated with the use of O3 was greater compared to the control group (53% vs 33%). However, there was no difference in hospital stay (9.37 ± 3.84 vs 9.37 ± 5.38; p=1).

There is a potential role for ozonated autohemotherapy for treatment of patients with severe COVID-19 pneumonia, with several biological plausible mechanisms of action. When human blood is exposed to a gas mixture of oxygen and ozone, oxygen equilibrates with the extracellular and intraerythrocytic water before becoming bound to hemoglobin until it is fully oxygenated. On the contrary, ozone, more soluble than oxygen, readily dissolves in water and reacts instantaneously with biomolecules, such as amino acids (particularly cysteine, tryptophan, methionine, phenylalanine, and tyrosine) and with lipids (particularly the unsaturated fatty acids contained in membrane phospholipids). The former can yield disulfides and methionine sulfoxide; the latter can yield hydrogen peroxide, aldehydes, and hydroxyhydroperoxides. The compounds generated during the reactions [reactive oxygen species (ROS) and lipid ozonation products (LOPs)] represent the “ozone messengers” and are responsible for its biological and therapeutic effects ^33^ so ozone can be considered as a pro-drug that produces biochemical messengers.

Regarding to the specific potential action of the ozone against coronavirus and the effectiveness of ozone against pathogens is well known. The ozone appears to be the best agent available for sterilizing water ^34^, although the in-vivo virucidal activity of ozone in the dosage used in this present study is unknown. It has been suggested that ozone could act a signal molecule in the organism, being generated by human neutrophils and being necessary for antibody-catalyzed formation ^35^ which play a role in the natural humoral response to infection. ^36^ Ozone also is capable of inducing the release and modulation of IFN-γ, TNF-α and colony stimulating factors, ^37, 38^ and is also able to modulate and stimulate phagocytic function ^39, 40^ which may have a very positive effect in COVID-19 infection.

Finally, ozone may impair viral replication, as suggested in its effects on SARS and MERS. ^41^ Angiotensin-converting enzyme type 2 (ACE2) cell receptors has been identified as receptor for SARS-CoV-2 ^42^, which could be blocked with specific monoclonal antibodies but also through the control of the nuclear factor erythroid 2–related factor 2 (Nrf2) that regulates and blocks the activity of this receptor. ^43^ Because ozone is able to cause a rapid Nrf2 activation, ^44, 45^ it seems very likely that this may be an important physiological mechanism to block endogenous COVID-19 reduplication by preventing contact with this receptor. Furthermore, spike proteins (S) is responsible for receptor binding and membrane fusion. ^46^ It contains a highly conserved transmembrane domain that consists of three parts: a N-terminal tryptophan-rich domain, a central domain, and a cysteine-rich C-terminal domain. Both, the cysteine-rich domain and tryptophan-rich domain, have been shown to be necessary for fusion. ^46-48^ Both cysteine and tryptophan, are sensitive to oxidation. It has been hypothesized that ozone metabolites could oxidize cysteine residues, making it difficult for the virus to enter the host cell and preventing viral replication. ^49^ This proof of concept study points to the need for further research, such as a well-designed, well-powered multicenter randomized clinical trial. Limitations include the sample size of our cohort is small and single-centered. The 95% CIs for our adjusted estimates were wide, and do not exclude a 20–30% decrease in the coefficient for time (days) to clinical improvement. Outcome assessors were not blinded to the treatment arm assignment. The group who received ozonated autohemotherapy were slightly younger and had lower body mass index. However, a post-hoc sensitivity analysis adjusted for age, quick SOFA and weight was conducted and the adjusted analysis confirmed the results. Furthermore, as it was an observational study IL-6 and other cytokines could not be measured. The strengths of this study include its pragmatic real-world COVID-19 population, use of objective primary clinical outcome and risk-adjustment using methods of regression modeling analyses.

In conclusion, ozonated autohemotherapy was associated with a significant shorter time to clinical improvement and shorter time to a two-fold decrease of C-reactive protein, ferritin, D-dimer and Lactate Dehydrogenase in severe COVID-19 pneumonia patients in this prospective case-control study.

## Data Availability

The authors confirm that the data supporting the findings of this study are available within the article [and/or] its supplementary materials.

## Acknowledgements

Dr. Vives affirm that has listed everyone who contributed significantly to the work.

Dr. Wijeysundera is supported in part by a Merit Award from the Department of Anesthesiology and Pain Medicine at the University of Toronto, and the Endowed Chair in Translational Anesthesiology Research at St. Michael’s Hospital and University of Toronto.

“This manuscript has been released as a pre-print at MedRxiv”. ^50^

## Notes

### Competing Interest Statement

The authors have declared no competing interest.

### Clinical Trial

NCT04444531

### Author Declarations

This study was conducted in compliance with the Declaration of Helsinki and approved by a multidisciplinary human research ethics committee at the institution.

